# SARS-CoV-2 & Pediatric Mental Health: A Review of Recent Evidence

**DOI:** 10.1101/2020.06.28.20136168

**Authors:** Mir Ibrahim Sajid, Javeria Tariq, Ayesha Akbar Waheed, Dur-e-Najaf, Samira Shabbir Balouch, Sajid Abaidullah

**Affiliations:** Student, Medical College, Aga Khan University, Stadium Road, Karachi; Assistant Professor, Oral & Maxillofacial Surgery, Neela Gumbad, Lahore, Pakistan; Professor of Medicine, King Edward Medical University, Neela Gumbad, Lahore, Pakistan; Head of North Medical Ward, Mayo Hospital, Neela Gumbad, Lahore, Pakistan

**Author notes:** Correspondence to: Mir Ibrahim Sajid, Medical College, Aga Khan University, Stadium Road, Karachi, Pakistan. 74880. Email Addresses (in order).

**Keywords:** SARS-CoV-2, COVID-19, Pediatrics, Mental Health, Stress, Anxiety, Depression

## Abstract

SARS-CoV-2 was declared a global pandemic by the World Health Organization (WHO) and was met with lockdown policies to curb the spread of the disease. This meant that 890 million students in 114 countries would be affected by the closure of their educational institutes, affecting their mental health. Mental health disorders are suggested to have a well-correlated link to suicide, which is the third most leading cause of death worldwide amongst children aged 15-19 years. According to WHO, “health is a state of complete physical, mental and social wellbeing and not merely the absence of disease.” Hence the isolation brought about by SARS-CoV-2 is postulated to cause anxiety, fear, and depression amongst the pediatric population, due to the loss of socialization and separation from friends. In this systematic review and meta-analysis, we highlight the major mental health issues in children aged 2-18 years, along with their causes, effects, and potential solutions to tackle these problems.

## INTRODUCTION

The novel coronavirus (n-CoV) stemmed as a local epidemic in the Wuhan City of China and presented as viral pneumonia of unknown aetiology. Once the nasopharyngeal samples were utilized to do a reverse transcriptase-polymerase chain reaction (RT-PCR) testing, it became evident, that this was a mutated version of the notorious Coronavirus family ^1^. What started as a Wuhan endemic disease, soon spread to other parts of the world and was declared as ‘pandemic’ by the World Health Organization on 12th March 2020 ^2^. To date, 16th June 2020, this virus infected more than 9 million people with >470,000 deaths (mortality rate: 5.19%).

In an attempt to reduce the spread of viral disease, lockdown policies were enforced all around the globe. This measure included the closure of the school and other educational institutes. According to an estimate from United Nations Educational, Scientific and Cultural Organization (UNESCO), 890 million students in 114 countries would be impacted due to these policies, which makes up for 86% of the world’s student population ^3^. Even though the closure of institutes in an attempt to curb a health crisis is not a new policy, however, the global exponential increase in the viral spread and disruptions of travel in terms of preventing students from reuniting with their family in such times ^4^, can and has caused significant psychological distress ^5^.

In this review, we highlight the major themes of mental health which have impacted the children during SARS-CoV-2. These children range from ages 2-18, and hence includes mostly school children and early collegiate students. We also discuss the challenges to the mental health of children who are already suffering from a particular disease process such as Cystic Fibrosis and Attention Deficit Hyperactivity Disorder.

## METHODS

This systematic review addresses the impact SARS-CoV-2 had on the pediatric population during SARS-CoV-2. This pediatric population would include individuals who are either tested positive for the viral infection or not and include individuals who were either healthy before this viral infection or were already going through a specific disease process. The PICO Question for this particular review is:

### Population

Pediatric population (age range: 2-18 years)

### Intervention

SARS-CoV-2

### Control

No disease process, healthy individuals, and individuals who were suffering from any disease other than SARS-CoV-2

### Outcomes

Mental health outcomes such as Stress, Anxiety, Anger and Depression

### Study Design

Case-control, cross-sectional, cohort studies and case series (cases reported >4), letters, perspectives and correspondences

### Time Frame

The disease duration: 2019-2020

### Search Strategy

An exhaustive literature review was conducted on four major databases, PubMed, CINAHL, Science Direct and Wiley Online Library; and a pre-print server, medRxiv to capture grey literature. The following search string was used: ((“COVID” OR “SARS” OR “n-CoV” OR “Novel Coronavirus”) AND (“Mental Health” OR “Depression” OR “Anxiety” OR “Stress” OR “Anger”) AND (“Child” OR “Paediatric” OR “Pediatric” OR “Adolescent”)). Concerning medRxiv, the search strategy was changed to multiple short searches using the keywords identified in the above search string. The following filters were used to narrow down the search: published in the years: 2019-2020, articles published in English Language and human subjects. A further filter used in Science Direct search included the article type to narrow down the search to only research articles and case reports.

### Selection Strategy

All the articles were downloaded in a new Endnote library, and two independent authors screened the non-duplicate articles on the base of title and abstract according to the following selection criteria. Any discrepancies in selection were resolved by a third independent author.

## Selection Criteria

### Inclusion Criteria

1. Addressing mental health issues pertaining to depression, anxiety, and stress
2. Articles addressing mental health issues during SARS-CoV-2
3. Articles addressing mental health in either healthy individuals or hospitalized patients.
4. Articles published in the English Language
5. Case-control studies, cross-sectional studies, cohort papers and case series (cases reported >4)
6. Letter to the editors and correspondences were included to increase the depth of review

### Exclusion Criteria

1. Articles addressing the parent’s conception of a child’s mental health
2. Review articles, case reports (cases reported<4), and clinical trial protocols

Initially, the authors decided to exclude letter to the editors, perspectives and correspondences, however, since most articles on this topic were based on such formats, and exclusion of these articles would decrease the content of the manuscript; the authors have decided to include them. The shortlisted articles were then downloaded for full-text review according to the inclusion criteria mentioned above, and any discrepancies in selection were resolved by a third independent author.

### Data Extraction & Analysis

The shortlisted articles were then extracted on a structured excel worksheet. Separate qualitative and quantitative analysis spreadsheets were developed to aid in the review process. Quantitative analysis was carried out using Cochrane’s Review Manager-aiding in meta-analysis and develop of the forest-plot diagram.

### Risk Bias Assessment

To carry out a risk bias assessment, Newcastle Ottawa Scoring (NOS) System was used for case-control study and modified NOS scale used for Cross-Sectional Study. Articles which did report bias were excluded from the library. NOS measures the risk bias and quality of the research on bases of three variables: selection, comparability and exposure. A score of 7 or more is expected to be a good score. Any study which reported a score of less than 7 was excluded from the final library.

## RESULTS & DISCUSSION

After a thorough screening process, a total of thirteen studies were shortlisted, which included one pre-print article from MedRxiv. PRISMA guidelines were followed to ensure standardization. A step by step record of included and excluded studies were recorded in a PRISMA flow diagram, displayed in Figure 1.

**Figure 1:**
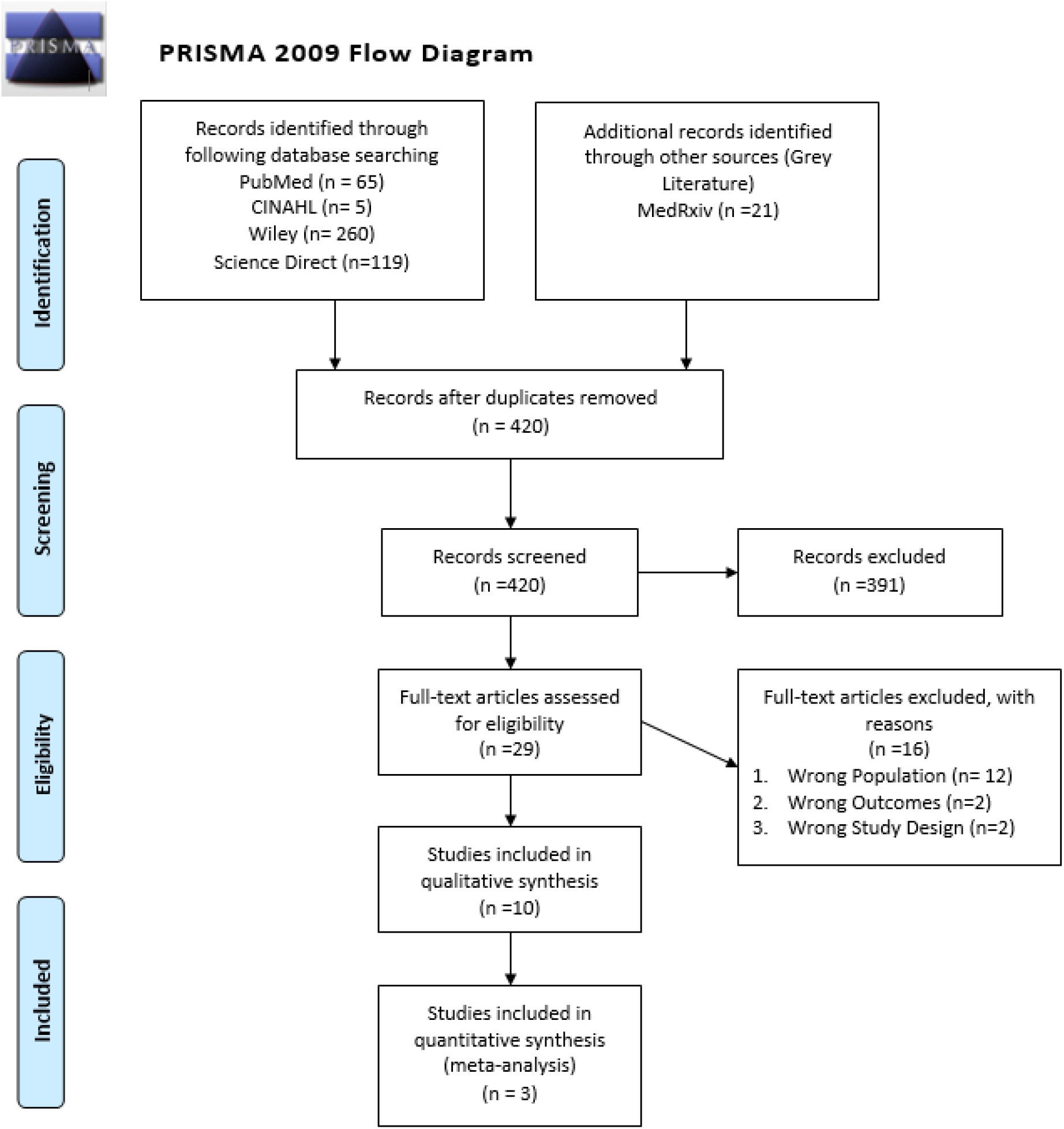
PRISMA Flow Diagram.

Risk bias assessment of all shortlisted studies was carried out using a NOS score or a modified NOS score as shown in Table 1. All four studies eligible for analysis had a score of ≥ 7 and hence were included in the study.

**Table 1:**

| **Table 1** |  |  |  |  |  |  |  |  |  |  |
| --- | --- | --- | --- | --- | --- | --- | --- | --- | --- | --- |
|  | **Selection** |  |  |  | **Comparability** |  | **Exposure** |  |  |  |
| **Study** | 1 | 2 | 3 | 4 | 1a | 1b | 1 | 2 | 3 | Total score |
| Senkalfa et al. | * | * | * | * | * | * | * | * | - | 8 |
| Shahidi SH et al. | * | * | * | * | * | - | * | * | - | 7 |
| Zhang J et al. | * | * | * | * | * | - | * | * | - | 7 |
| Zheng Y et al. | * | * | * | * | * | - | * | * | * | 8 |

Table 2 shows the detailed characteristics of the studies included for review. As explained in the methods, to expand the pool of knowledge, the authors had to include qualitative accounts such as editorials and viewpoints. Only two studies involved children who were already involved in a disease process, i.e. Senkalfa et al. who evaluated the mental health status of patients suffering from cystic fibrosis and Zhang J et al. who explored SARS-CoV-2 mental health impact on patients with ADHD.

**Table 2:** Characteristic of Selected Stdies

| **Table 2: Characteristic of Selected Studies** |  |  |  |  |  |
| --- | --- | --- | --- | --- | --- |
| **Author** | **Study Title** | **Journal** | **Study Design** | **Study Population** | **Risk Bias Assessment (NOS)** |
| Dalton L et al. | Protecting the psychological health of children through effective communication about COVID-19 | Lancet Child Adolescent Health | Viewpoint | Children | - |
| Green P | Risks to children and young people during covid-19 pandemic | BMJ | Editorial | Children | - |
| Lee J | Mental health effects of school closures during COVID-19 | Lancet Child Adolescent Health | Viewpoint | School Children | - |
| Liu JJ et al. | Mental health considerations for children quarantined because of COVID-19 | Lancet Child Adolescent Health | Viewpoint | Children who are quarantined | - |
| Novins DK et al. | JAACAP's Role in Advancing the Science of Pediatric Mental Health and Promoting the Care of Youth and Families During the COVID-19 Pandemic | Journal of American academy of child and adolescent psychiatry | Editorial | Children, adolescents, and their families | - |
| Rahman M et al. | Risks to Bangladeshi children and young people during covid-19 outbreak | BMJ | Editorial | Children and young people | - |
| Senkalfa et al. | Effect of the COVID-19 pandemic on anxiety among children with cystic fibrosis and their mothers | Pediatric Pulmonology | Case-control | Children aged 0-18 with Cystic Fibrosis and their mothers | 8 |
| Shahidi SH et al. | Physical activity during COVID-19 quarantine | Acta Paediatrica | Case-control | Children | 7 |
| Walters A | Supporting youth and families during COVID-19 | The Brown University Child and Adolescent Behaviour | Editorial | Pre-school, School-age children and adolescents | - |
| Wang G et al. | Mitigate the effects of home confinement on children during the COVID-19 outbreak | Lancet Child Adolescent Health | Article | Children | - |
| Zhai Y et al. | Addressing collegiate mental health amid COVID-19 pandemic | Psychiatry Research | Editorial | College students | - |
| Zhang J et al. | Acute stress, behavioral symptoms and mood states among school-age children with attention-deficit/hyperactive disorder during the COVID-19 outbreak | Asian Journal of Psychiatry | Cross-sectional survey | ADHD Children | 7 |
| Zheng Y et al. | A survey of the psychological status of primary school students who were quarantined at home during the coronavirus disease 2019 epidemic in Hangzhou | MedRxiv | Cross-sectional survey | Students from grade 1 to grade 6 enrolled in two primary schools (one private and one public) in the urban area of Hangzhou, China | 8 |

The authors then discussed the major mental health themes identified in these papers. Among the major mental health issues identified were stress, anxiety, depression, boredom and the fear of getting infected. Table 4 discusses all of these major themes in detail and provides a brief explanation for all of these findings. The table also briefs on possible solutions identified in the paper regarding the problems highlighted in the paper.

**Table 3:** Major Mental Health Themes & Solution Identified

| Author | Major Mental Health Issue Identified | Brief Explanation for Observations Noted | Solutions Discussed |
| --- | --- | --- | --- |
| Dalton L et al. <sup>6</sup> | Stress<br>Anxiety | Children are exposed to and aware of stressful information and cues from the behaviour of adults around them, causing anxiety.<br>Children struggle to make sense of the changes around them when they are deprived of the acknowledgement and information of the problems, they see around them.<br>Children might see these difficult times, such as illness and loss, as a punishment for their behaviour.<br>Adults may be desensitized to their children's psychological needs owing to the stress of SARS-CoV-2 itself.<br>Children may remain silent or exhibit unusual behaviour (e.g. anger, tears, sadness) | Honest, open-ended communication regarding the situation from adults, allows children to open up and manage stress and anxiety better.<br>Adults should not overwhelm children with their fears.<br>This communication should take into account an assessment of the child's depth of understanding.<br>To find out how children are feeling, adults must first be the role models of sharing feelings.<br>Children are then allowed closure for what they observe and open up. |
| Green P <sup>7</sup> | Anxiety<br>Abuse | In certain family settings, prolonged lockdown can lead to an atmosphere that leaves children vulnerable to abuse and neglect. | A top to bottom approach at the national and governmental level can address these dangers, more so after |
|  |  | • Simultaneously, there is a lack of 'safeguarding' from outside sources such as teachers, health workers, and social workers, who can identify patterns of abuse.<br>• SARS-CoV-2 has produced patterns that have led to a rise in police involvement in domestic abuse cases, child support line calls. | the immediate danger of the pandemic subsides.<br>• Talks by leaders and community alike should address the needs of children during this time.<br>• This global event should become a precipitating factor for reform. |
| Lee J<sup>8</sup> | • Stress<br>• Anxiety<br>• Depression<br>• Special needs | • Both otherwise healthy children and those facing depression lack a structured routine because of the suspension of educational activity.<br>• The lack of structure to the day has induced anxiety and depression in some and worsened that of those already suffering from the above-mentioned diseases.<br>• Children with special needs are even more predisposed to distress and anxiety because of the sudden shift in lifestyle.<br>• Postponing of major life events combined with the stress to 'stay healthy' for multiple age groups, including college students, has led to an increase in stress levels. | • As not enough is known about the nature of this phenomenon, there is a need to conduct more research to develop the competency to address the issue.<br>• Development of creative ways to substitute for social support groups and counselling, such as shifting them online<br>• Monitor and follow-up with the mental health status of these age groups long term as a means of investigation. |
|  |  | • Social isolation in abusive homes in a climate that has increased the incidence of abuse itself can further contribute to mental health issues. |  |
| Liu JJ et al.<sup>9</sup> | • Stress<br>• Anxiety<br>• Post-traumatic stress disorder | • Children who are separated from their caregivers either because they have the infection themselves or because their caregiver has contracted it are at the greatest risk for developing mental health disorders<br>• These include acute stress disorder, post-traumatic stress disorder and depression.<br>• Separation and loss of parents at this stage translate to long term mental health problems. | • Nurses and other support systems are made available 24/7 for children in isolation wards in China's largest cities.<br>• Children are made to use electronic devices to communicate with parents.<br>• Volunteers act as caregivers for children who have recovered and cannot be united with their parents.<br>• Pediatric healthcare workers to receive formal training to identify children's mental health problems at the initial stages.<br>• Producing evidence-based guidelines along with operational protocols during SARS-CoV-2 to streamline the process |
|  |  |  | • Improving access to mental health services for children<br>• Post-Pandemic surveillance |
| Novins DK et al. <sup>10</sup> | All general mental health disorders | • The article highlights areas of interest that are to be investigated, including the impact of the pandemic on children's mental health.<br>• Outlines different ways of assessing impact, including but not limited to novel research, genetic studies, clinical knowledge | • Conducting research will provide a database for decision making.<br>• Research on the effectiveness of support systems and interventions addressing mental health<br>• Mentions investigations of preemptive and preventive measures, high-risk groups and outcomes of protocols implemented. |
| Rahman M et al. <sup>11</sup> | • Anxiety<br>• Post-traumatic stress disorder | • A sedentary lifestyle contributes to obesity and other chronic illnesses<br>• Domestic abuse rates have increased during the conditions produced by the lockdown.<br>• Compounding factors such as parental anxiety, loss of income, and uncertainty about SARS-CoV-2 can lead to mental health disorders such as PTSD. | • Initiation of 'community-based' programs and protocols to address the need for reduction in these poor outcomes. |
|  |  | The sample population of Bangladesh face higher levels of undernutrition, as well as adverse circumstances conditions such as bondage to child labour and homelessness. They are especially susceptible to mental health decline. |  |
| Senkalfa et al. <sup>12</sup> | Anxiety<br>Depression | The impact of CF during SARS-CoV-2 on the anxiety levels of children and their mothers- both healthy and affected were analyzed according to STAI<br>Mothers of children with CF had higher anxiety levels<br>Healthier children had higher anxiety than those affected<br>Tele-conferencing for the interview reduced mothers' anxiety about SARS-CoV-2<br>The control group faced higher levels of anxiety potentially because they have not faced the fear and burden of disease like the study group | Consulting a physician who is known to families about the unprecedented situation and offering an interview will bring relief.<br>Tele-conferencing with parents and families of children with chronic illness can address fears and reduce anxiety. |
| Shahidi SH et al. <sup>13</sup> | Lack of physical activity influences cognitive, emotional and social components of development. | Physical literacy tends to enhance confidence and improves motor skills as demonstrated by children receiving school-based circus arts instructions when compared to ones | Implementation of online programs to keep children mentally engaged indoors |
|  |  | receiving standard physical education. | ● Restriction on-screen time to prevent inactivity |
| Walters A <sup>14</sup> | ● Preschool children: Increased clinginess, bedwetting, irritability, disobedience, fear<br>● School-age children: anxiety, worry, loss of typical activity<br>● Adults: isolation, substance abuse, decreased responsiveness. | Withdrawal of economic and emotional support in difficult times have precipitated existing mental health issues and given rise to new. | ● Use of counselling therapies<br>● Interventions delivered in less than 4 months<br>● Interventions with the involvement of family |
| Wang G et al. <sup>15</sup> | ● Increased stress, frustration, boredom<br>● Fear of confinement and getting infected | Mean post-traumatic stress scores appeared to be 4 times higher in quarantined children when compared to non-quarantined ones. | ● The government must create awareness of the consequences<br>● The government should predefine effective online learning standards<br>● Online service of a psychologist<br>● Schools, NGOs, social workers must offer counselling for children<br>● Good parenting skills |
|  |  |  | • Videos for SARS-CoV-2 precautions |
| Zhai Y et al. <sup>16</sup> | • Anxiety<br>• Depression<br>• Substance abuse<br>• Sleeping disorder<br>• Eating disorders | Universities were not able to fully administer college students' health, education and safety needs. | • Provision of remote education<br>• Alternatives for internships and research projects halted |
| Zhang J et al. <sup>17</sup> | • Worsening of symptoms of Attention Deficit Hyperactivity Disorder (ADHD) in children during SARS-CoV-2 outbreak | One sample t-test revealed deteriorated average ADHD behaviour during SARS-CoV-2 outbreak when compared to the normal state | • Online education<br>• Increased online study hours |
| Zheng Y et al. <sup>18</sup> | • Increased anxiety in primary school students quarantined at home. | Mean total score SASC (Social Anxiety Scale for Children) was higher in quarantine primary school children in comparison with children from urban areas. | • Early identification and intervention of mental health issues in quarantine children<br>• Guidance from parents and family members to children<br>• Scientific education about SARS-CoV-2 to decrease fear and anxiety<br>• Maintaining the regular schedule before being quarantine. |

**Table 4:** Characteristic of Quantitative Studies

| **Author** | **Mean Age of Study Population** | **Gender Ratio (M: F)** | **Mean Age of Control Group** | **Gender Ratio of Control Group** | **Mental Health Concerns In Study Population** | **Mental Health Concern In Control Group** |
| --- | --- | --- | --- | --- | --- | --- |
| Senkalfa et al. | • 25 patients were aged 0–8 years<br>• 14 patients were aged 9–12 years<br>• 6 were aged 13–18 years | 5.1:4.9 | • 46 patients were aged 0–8 years<br>• 22 patients were aged 9–12 years<br>• 22 patients were aged 13–18 years | 5.1:4.9 | • Children with Cystic fibrosis had no elevated anxiety levels since they suffer constant diseased anxiety.<br>• State anxiety for age groups 9–12 was almost the same for both groups ($28.4 \pm 5.2$ in study population vs $29.4 \pm 4.1$ in the control group) with a p-value of 0.547<br>• Trait anxiety scores for age groups 9–12 was not significantly different in both groups ($35.2 \pm 7.6$ in study group vs $31.9 \pm 5.7$ in the control group) with a p-value of 0.172<br>• Neither state, not trait anxiety scores differed | • State anxiety scores for children aged 13–18 years in the control group were higher than for the age-matched children in study population ($29.0 [27.8-32.3]$ vs $41.5 [35.5-46.3]$; $p = 0.003$)<br>• Trait anxiety scores for children aged 13–18 years in the control group were not significantly different than for the age-matched children in the study population [$34 (30.8-36.5)$ in study group vs $39 (32-42.3)$ in the control group]<br>• Mothers on teleconference; 34.4% reported a decrease in anxiety, while 12.2% reported an increase, |
|  |  |  |  |  | by gender for 9–12-year-olds and 13–18-year-olds in both study and control groups ($p > 0.05$).<br>Informing parents of children with CF about SARS-CoV-2 by teleconference may decrease anxiety. (84% reported a decrease in anxiety, while 6.7% reported an increase, and 8.9% reported no change) | and 53.3% reported no change.<br>Increased fear and anxiety of facing a serious health risk for the first time due to pandemic. |
| Zhang J et al. | $9.43 \pm 2.39$ | 8.05:1.95 | The same population before the pandemic | The same population before the pandemic | Children's ADHD behaviour worsened during the pandemic as compared to their normal state as illustrated by a higher one sample t-test score of $2.25 \pm 0.54$<br>Ability to stay focused, listen to instructions, and keep routine got worse<br>Anger frequency severely worsened<br>Limited access to psychiatric medicine during the pandemic | Ability to keep the room neat, to work quietly, to keep the adults uninterrupted, eating and sleeping behaviour showed no significant change (approximately 50% or greater children did not show much difference) |
| Zheng Y et al. | 10.10 ± 1.63 | 5.22:4.78 | - | - | • Higher level of anxiety as depicted by a greater mean SASC (Social Anxiety Scale for Children) score of $3.90 \pm 3.73$ compared to a lower score of control group $3.48 \pm 3.47$<br>• Female students had significantly higher SASC scores than the control group scores ($P < 0.01$) on all three measures (fear of negative evaluation, social avoidance and distress)<br>• Male students had significantly higher scores than the control group only for fear of negative evaluation ($P < 0.01$)<br>• Risk factors for anxiety:<br>1) being male<br>2) Children accompanied by Fewer relatives ($P < 0.05$)<br>3) deterioration of the parent-child | • Higher level of depression as depicted by a greater mean DSRSC (Depression Self-rating Scale for Children) score of $(9.84 \pm 4.73)$ compared to a lower score of study group $5.67 \pm 4.97$ Hence ($P < 0.05$)<br>• Risk factors for depression:<br>1) older age (mean DSRSC score of the high-grade group (grades 4-6) was $6.04 \pm 5.28$, which was significantly higher than those of the low-grade group (grades 1-3) ($5.06 \pm 4.33$) ($P < 0.01$))<br>2) a poor parent-child relationship (3.697 times more likely to suffer)<br>3) deterioration of the parent-child relationship (5.744 times more likely to suffer)<br>4) less physical activity |
|  |  |  |  |  | relationship($P < 0.05$) (2.068 times more likely to suffer)<br>4) increased conflicts with parents<br>5) irregular learning and rest<br>6) worrying about being infected (2.206 times more likely to suffer)<br>7) negative mood | 5) irregular work and rest<br>6) negative mood (2.438 times more likely) |

Three papers were identified to be included in the quantitative analysis; Senkalfa et al., Zheng Y et al., and Zhang J et al. A detailed highlight of data extracted from these studies can be visualized in Table 5. We address the major mental health determinants in the study population and pin them against the control population using means and standard deviations for the ease of comparison.

**Table 5:** Conclusions

| Table 5: Conclusions |  |
| --- | --- |
| Major Health Concerns Amongst Pediatric Population during SARS-CoV2 pandemic | Potential solutions to counter the deteriorating mental health status |
| • Anxiety<br>• Stress<br>• Boredom<br>• Fear of getting infected<br>• Feeling of loneliness or confinement<br>• Grief/sorrow<br>• Anger and disobedience<br>• Negative emotions and thoughts<br>• Depression<br>• Sleeping and eating disorders<br>• Worsening of existing health condition such as ADHD or cystic fibrosis<br>• Fall in academic performance<br>• Conflicts with parents and siblings | • Physical activity/exercise at home helps release hormones that enlighten ones mood<br>• Parents to promote a health home environment ensuring quality family time<br>• Getting oneself enrolled in online courses and activities of interest<br>• Video calling to allow a virtual get-together<br>• A fixed routine with a proper to-do schedule |

A statistical analysis of whether SARS-CoV-2 has impacted mental health has been carried out on two papers eligible for a meta-analysis. The papers authored by Senkalfa et al. and Zheng et al. analyzed the development of anxiety between the study and control population. Cochrane’s Review Manager was utilized to carry out the meta-analysis and make a forest plot, as shown in Figure 2. The analysis revealed an overall z-score of 0.85 (p=0.39) with Heterogeneity: Tau^2^ = 3.27; Chi^2^ =48.44, df= 1 (p<0.00001); I^2^ = 98%. It is evident that there exists significant heterogeneity in the studies and that there was a non-significant weak correlation between anxiety in the case and control group.

**Figure 2:**
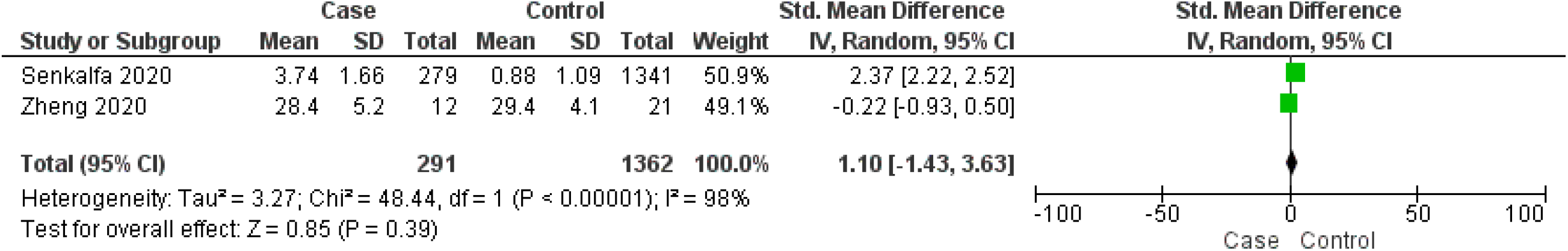
Forest-Plot Diagram.

A global pandemic such as SARS-CoV-2 can bring with it, a huge toll on an individual’s psychology. Past research and recent data ^19^ have shown that individuals can exhibit fear and anxiety-prone distress responses which can either stem from a fear of getting infected themselves, or of their loved ones, and the socio-economic consequences this crisis can bring with it- such as loss of jobs. The loss of jobs can directly impact the growth and development of the child, directly impacting their mental health. Low and Middle-Income Countries (LMIC) such as Pakistan have a large majority of the population who are either daily wagers or depend on their weekly incomes to feed their families. With predicted three million people losing jobs (one million in industrial; two million in the services sector) and poverty headcount to soar from 24.3% to 33.5% ^20^- many families are expected to be pushed towards starvation, rendering children more vulnerable to an already prevailing malnutrition and mental health deterioration.

It is of vital importance to make sure that the time children spend at home during the lockdowns does not negatively impact their mental health. Among things which should be taken into consideration, the excessive use of the internet cannot be ignored. Deslandes and colleague ^21^ pointed out how excessive use of the internet during home isolation can render an individual prone to self-harm. This observation comes amidst fear of the ‘unseen virus’ and the easily available paranoia of infections and death tolls in their communities. Hence, organizations such as WHO and UNICEF ^22, 23^ suggest that the information on the pandemic must be controlled by the parents, without hiding facts, however ensuring that it is communicated in a medium that can be easily understood by children of different age groups. This will enable them to validate their emotions and express their feelings, about the ongoing circumstances.

## CONCLUSION

An analysis of available literature illustrates a clear correlation between increasing anxiety amongst children during the SARS-CoV-2 pandemic. Our systematic review encompasses school-going and early collegiate students who have shown to suffer from a deteriorating mental status due to the fear of being infected, a feeling of boredom, and lack of productivity, which adds on to the stress level hindering their focused approach to learning. A quick summary of our review is made in Table 5.

## Data Availability

All the data included in the paper has been added in the manuscript and in freely available.

## DISCLOSURES

### Ethics approval and consent to participate

All procedures performed on patient laboratory samples in this study were in accordance with the ethical standards of the Institute Ethics Committee and with the 1964 Helsinki declaration and its later amendments or comparable ethical standards.

### Consent for publication

Not Applicable

### Availability of data and materials

The corresponding authors would be highly obliged to share the relevant data and materials of this work on the request of the journal’s editor.

### Author Contributions

Mir Ibrahim Sajid and Javeria Tariq were involved in manuscript writing and analysis, Ayesha Akbar Waheed and Dur-e Najaf were involved in the screening of the papers using title, abstract and later full text, Samira Shabbir Balouch and Sajid Abaidullah supervised the project and proof-read before submission.

## Acknowledgements

None

## REFERENCES

1. C. Huang YW, X. Li, L. Ren, J. Zhao, Y. Hu, L. Zhang, G. Fan, J. Xu, X. Gu, Z. Cheng, T. Yu, J. Xia, Y. Wei, W. Wu, X. Xie, W. Yin, H. Li, M. Liu, Y. Xiao, H. Gao, L. Guo, J. Xie, G. Wang, R. Jiang, Z. Gao, Q. Jin, J. Wang, B. Cao. Clinical features of patients infected with 2019 novel coronavirus in Wuhan, China. Lancet (London, England) 2020; 395(10223): 497–506.

2. Sajid M, Awais S, Salar K, et al. Impact of COVID-19 on pregnancy and childbirth: a systematic review of recent evidence. Biomedica 2020; 36: 145–57.

3. UNESCO. COVID-19 educational disruption and response. Available at: https://en.unesco.org/themes/education-emergencies/coronavirus-school-closures. 2020.

4. Zhai Y, Du X. Mental health care for international Chinese students affected by the COVID-19 outbreak. Lancet Psychiatry 2020; 7(4): e22.

5. Araújo FJO, de Lima LSA, Cidade PIM, Nobre CB, Neto MLR. Impact Of Sars-Cov-2 And Its Reverberation In Global Higher Education And Mental Health. Psychiatry Res 2020; 288: 112977.

6. Dalton L, Rapa E, Stein A. Protecting the psychological health of children through effective communication about COVID-19. The Lancet Child & adolescent health 2020; 4(5): 346–7.

7. Green p. Risks to children and young people during covid-19 pandemic. BMJ 2020; 369: 1669.

8. Lee J. Mental health effects of school closures during COVID-19. Lancet Child Adolesc Health 2020; 4(6): 421.

9. Liu JJ, Bao Y, Huang X, Shi J, Lu L. Mental health considerations for children quarantined because of COVID-19. Lancet Child Adolesc Health 2020; 4(5): 347–9.

10. Novins DK, Althoff RR, Billingsley MK, et al. JAACAP’s Role in Advancing the Science of Pediatric Mental Health and Promoting the Care of Youth and Families During the COVID-19 Pandemic. J Am Acad Child Adolesc Psychiatry 2020; 59(6): 686–8.

11. Rahman MS, Lassi ZS, Shariful Islam SM. Risks to Bangladeshi children and young people during covid-19 outbreak. BMJ 2020; 369:m2299.(doi).

12. Senkalfa BP, Eyuboglu TS, Aslan AT, et al. Effect of the COVID-19 pandemic on anxiety among children with cystic fibrosis and their mothers. Pediatr Pulmonol 2020.

13. Shahidi SH, Stewart Williams J, Hassani F. Physical activity during COVID-19 quarantine. Acta Paediatrica 2020.

14. Walters A. Supporting youth and families during COVID-19. The Brown University Child and Adolescent Behavior Letter 2020; 36(6): 8.

15. Wang G, Zhang Y, Zhao J, Zhang J, Jiang F. Mitigate the effects of home confinement on children during the COVID-19 outbreak. Lancet 2020; 395(10228): 945–7.

16. Zhai Y, Du X. Addressing collegiate mental health amid COVID-19 pandemic. Psychiatry Res 2020; 288: 113003.

17. Zhang J, Shuai L, Yu H, et al. Acute stress, behavioural symptoms and mood states among school-age children with attention-deficit/hyperactive disorder during the COVID-19 outbreak. Asian J Psychiatr 2020; 51: 102077-.

18. Zheng Y, Li J, Zhang M, et al. A survey of the psychological status of primary school students who were quarantined at home during the coronavirus disease 2019 epidemic in Hangzhou China. medRxiv 2020.

19. Taylor S, Landry CA, Paluszek MM, Fergus TA, McKay D, Asmundson GJG. Development and initial validation of the COVID Stress Scales. J Anxiety Disord 2020; 72: 102232.

20. Reuters. Pakistan’s deficit and poverty rate to soar due to coronavirus, govt estimates show. Dawn.com. 2020. Available from: https://www.dawn.com/news/1557111. 2020.

21. Deslandes SF, Coutinho T. The intensive use of the internet by children and adolescents in the context of COVID-19 and the risks for self-inflicted violence. Cien Saude Colet 2020; 25(1): 2479–86.

22. World Health Organization (WHO) U, Children EVA, Things IG, Health PfL, USAID, Center for Disease Control and Prevention, GCFL. COVID-19 parenting. Available at: https://www.covid19parenting.com. 2020.

23. UNICEF. How teenagers can protect their mental health during coronavirus. Available at: https://www.unicef.org/coronavirus/how-teenagerscan-protect-their-mental-health-during-coronavirus-covid-19. 2020.

